# Gender Differences in Caries and Periodontal Status in UK Children

**DOI:** 10.1101/2021.03.24.21253842

**Authors:** Sofia Papadaki, Gail V A Douglas, Alaa HaniBani, Jing Kang

**Author notes:** CORRESPONDING AUTHOR: Jing Kang. Author contribution statement: All authors substantially contributed to the conception and design of this study, drafting and revising the article critically, and final approved the version. SP and JK particularly performed the data acquisition, analysis, and interpretation. All authors declare no conflict of interest.

## Abstract

**Background:** Gender inequalities in dental caries and periodontal diseases have been observed among adults. However, literature is scarce for children and evidence on gender inequalities regarding caries and/or periodontal diseases is vague. Our aim is to examine potential gender differences in UK children regarding caries experience and periodontal status using data from the UK’s 2013 Children’s Dental Health Survey (CDHS).

**Methods:** CDHS included children aged 5, 8, 12 and 15 years. Their dental caries experience and periodontal status were reported using the number of decayed, missing and filled teeth (DMFT or dmft for permanent or primary dentition at both D1 and D3 thresholds) and the basic periodontal examination (BPE) score, respectively. Zero-inflated negative binomial (ZINB) models were used to fit DMFT/dmft and a multinomial logistic regression (MLR) model was used for BPE scores after adjustment for possible confounding factors, to assess the gender inequality on DMFT/dmft and BPE in the UK children.

**Results:** The analyses included 9,866 children. No gender inequalities in caries experience were observed in the 5 and 8-year-old children regardless of the threshold at which dental caries were examined. However, for the 12- and 15-year-old adolescents, females had higher D_3_MFT scores compared to males (IRR: 1.28, 95% CI: 1.10-1.49 and IRR: 1.16, 95% CI: 1.00-1.35, respectively). Additionally, the 15-year-old females had lower probability to be caries free (OR: 0.59, 95% CI: 0.45-0.82), regardless of the threshold at which dental caries were examined. With regards to the periodontal status, no statistically significant gender inequalities (p>0.05) were observed.

**Conclusions:** In the UK, female adolescents had experienced more carious lesions compared to males of the same age group, when dental caries were examined into dentine (D_3_MFT). However, 15-year-old males matched females in their caries experience, when the early enamel lesions were included in caries diagnosis (D_1_MFT). With regard to the periodontal status, no gender dissimilarity was confirmed among British adolescents. The increased risk of adolescent females to dental caries may signify additional needs for prevention and improved oral care.

## Introduction

Gender inequalities in dental caries and periodontal diseases among adults have been investigated widely. Many studies showed that females bear a higher burden of dental caries^1^, while males were found to be more susceptible to develop advanced periodontal disease^2^. However, evidence in the field of paediatric dentistry remains rather unclear, as conclusions from various studies are mixed and contradictory. Some published literature depicted gender bias regarding caries experience for juvenile patients, with females being reported as more susceptible^3-8^. On the other hand, there are studies supporting an opposite trend, indicating that males, especially at the primary dentition stage, exhibit higher caries experience than females of the same age ^9-12^. Moreover, some studies found no significant (p>0.05) gender inequalities in caries experience^13-17^.

For periodontal diseases, most literature suggests that male teenagers are more likely to develop gingivitis and have suboptimal oral hygiene^18-21^. However, some studies report the absence of significant gender inequalities in the periodontal status of children^22-24^.

Evidence around gender inequalities is as yet incomplete, with limitations including biased sampling, small sample size and methodological flaws. Therefore, this study aims to add information by examining potential differences in caries experience and periodontal status between genders using the UK 2013 Children’s Dental Health Survey (CDHS) data, a large and robust epidemiological survey. It is noteworthy that the CDHS is the first survey in the UK that measures caries at the D1 threshold. This novelty makes the current study interesting in that both enamel and dentine caries were measured. An affirmation of gender inequalities in children’s oral health is important, as this would stimulate further studies in order for the real cause of this assumption to be identified and therefore addressed.

## Material and methods

### Data source

The 2013 CDHS was used as the data source, including children from England, Wales and Northern Ireland. Data were collected through clinical examinations by calibrated dental teams, questionnaires completed by the 12- and 15-year-old pupils and parent questionnaires of those children who participated in the study^25^. The target populations were the 5-, 8-, 12- and 15-year-old children, representing the primary (5-year-old), mixed (8-year-old) and permanent dentition (12- and 15-year-old)^25^.

Ethical approval for CDHS was gained by the University ethics committee at the University College London (Project ID 2000/003) and participants had consented in advance for the use of data in future research projects.

### Outcome measures

There were two dental outcome measures:

1. Children’s dental caries experience based on DMFT/dmft scores
2. Periodontal status using the available BPE scores

Children’s dental caries experience was measured at two different thresholds: including enamel lesions (D1) and lesions only into dentine (D3). The CDHS was novel in measuring caries at the D1 threshold. Table 1 and Figure 1 demonstrates the differences of these two caries thresholds.

**Table 1.** Explanation of differences between D_1_MFT/d_1_mft (enamel lesions) and D_3_MFT/d_3_mft (dentine lesions).

| D<sub>1</sub>MFT/d<sub>1</sub>mft | D<sub>3</sub>MFT/d<sub>3</sub>mft |
| --- | --- |
| Decayed teeth including early enamel lesions along with dentinal caries | Decayed teeth, including caries lesions into dentine only |
| Missing teeth due to caries | Missing teeth due to caries |
| Filled teeth | Filled teeth |

**Figure 1.**
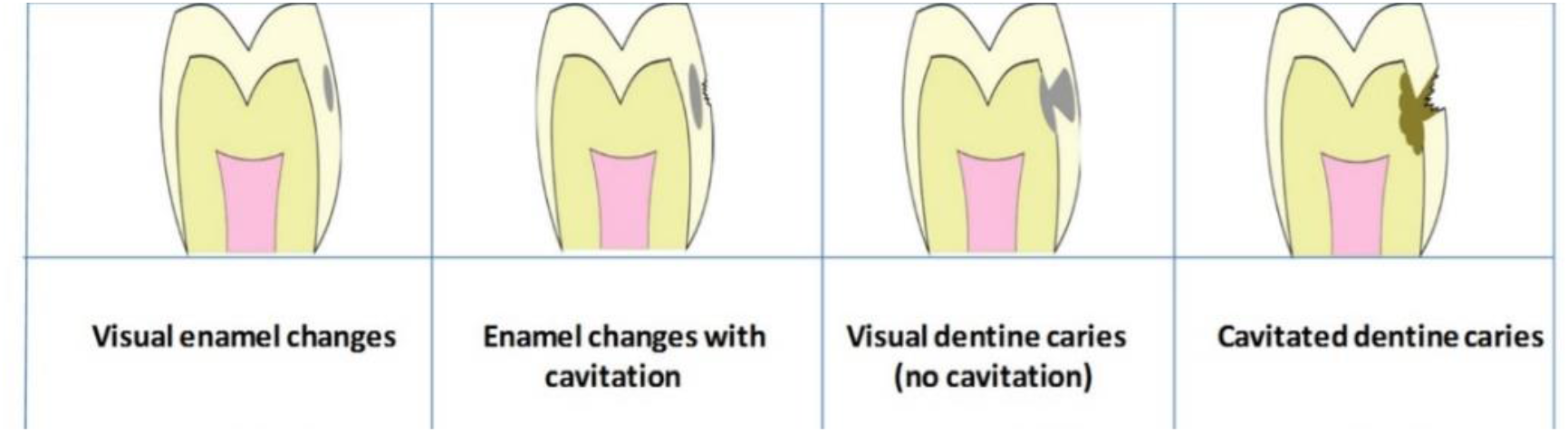
Illustration of the differences between D_1_MFT/d_1_mft (including enamel and dentine lesions) and D_3_MFT/d_3_mft (dentine lesions only)^49^.

The periodontal status was measured based on BPE scores (Table 2), which were only available for the 15-year-old children.

**Table 2.** BPE scores and levels in multiple regression analysis.

| Levels | BPE scores |  |
| --- | --- | --- |
| Level 1 | 0 | No bleeding |
| Level 2 | 1 | Bleeding on probing |
| Level 3 | 2 | Bleeding with retentive factors, such as calculus |
| Level 4 | 3 | Bleeding with shallow pockets between 3.5-5.5mm |
|  | 4 | Deep pockets>5.5mm |

### Exposure

Gender was the main exposure measure.

### Covariates

Covariates included participants’ demographic characteristics such as age (scale) and socio-economic status (SES; quantiles 1-5)^26^, their dietary habits such as consumption of water or food and drinks containing sugar including cakes, biscuits, sweets, fruit juice, smoothies, energy and fizzy drinks (less than once a day, more than twice a day), their oral hygiene practices such as tooth brushing frequency (less than once a day, more than twice a day), tooth flossing (yes, no) and pattern of dental attendance patterns^27^ (only when have problems arise, routinely), experience of orthodontic treatment (yes, no), as well as dental anxiety levels (low, medium, high)^28,29^.

More covariates were also examined, such as children’s experience of prevention (fissure sealants, fluoride varnish) and use of fluoride toothpaste. However, those covariates that were available for only a small number of participants or did not reach the set significance level at 0.5^30^ were not included in the multivariable analysis for the adjustment of the ZINB and MLR models.

### Statistical Analysis

Descriptive statistics was performed for comparisons between males and females. The zero-inflated negative binomial models (ZINB) were used to fit the D_1_MFT/d_1_mft and D_3_MFT/d_3_mft scores, as it was indicated by the DMFT/dmft distribution, with more than half of children having a DMFT/dmft score equal to zero. ZINB model is a two-part model, with the zero-inflated part estimating the chance of excessive zeros, and the count model the number of DMFT/dmft. The confidence interval (CI) was set to 95% after sampling weight. Adjustments for demographic features, oral hygiene practices, dietary habits and dental anxiety were included in the models gradually. This led to three different statistical models from a univariate to a multivariate approach to demonstrate the robustness of the results.

The m_0_ model was unadjusted comparing the D_1_MFT/d_1_mft and D_3_MFT/d_3_mft differences between genders without taking into consideration other possible confounders.

The m_1_ model was adjusted for demographic features.

The m_2_, which applied only for the D_1_MFT and D_3_MFT variables, was adjusted for the above-mentioned covariates, including children’s SES, dental behaviour, dietary habits and dental anxiety.

This happened as a result of limited information about the 5- and the 8-year-old children with regards to their oral hygiene and dietary habits.

Additionally, subgroup statistical analysis to estimate children’s caries experience in each age group was performed. The subgroup analysis referred to the 5-, 8-, 12- and 15-year-old children, respectively.

With regards to periodontal status, data were analysed using the Multinomial Logistic Regression (MLR) model at four different levels [level 1, or reference level (BPE:0), level 2 (BPE:1), level 3 (BPE:2) and level 4 (BPE:3 or 4)], with the aim to compare each group (levels 2, 3, 4) with reference level (Table 2). The model was sequentially adjusted for SES, oral hygiene practices, including frequency of toothbrushing, flossing, dental attendance pattern, and experience of orthodontic treatment^31^.

Data were weighed according to primary sampling unit (PSU) and strata, and then, analysis was performed using R studio with Survey Package and StataSE 14.

## Results

In total, 9,866 children were included in the analyses. A comparison of different covariates between genders is presented in Table 3.

**Table 3.** Comparison between male and female participants in 2013 CDHS. N=9866.

|  | Male | Female | p-value |
| --- | --- | --- | --- |
| N | 4812 (48.8) | 5054 (51.2) |  |
| **<u>Demographics</u>** |  |  |  |
| Age group |  |  | 0.496 |
| 5-year-old | 1264 (26.3) | 1285 (25.4) |  |
| 8-year-old | 1171 (24.3) | 1196 (23.7) |  |
| 12-year-old | 1222 (25.4) | 1310 (25.9) |  |
| 15-year-old | 1155 (24.0) | 1263 (25.0) |  |
| HMRC quintile |  |  | 0.001 |
| 1 (Most deprived) | 1432 (30.9) | 1669 (34.5) |  |
| 2 | 1062 (22.9) | 1144 (23.7) |  |
| 3 | 952 (20.6) | 903 (18.7) |  |
| 4 | 710 (15.3) | 671 (13.9) |  |
| 5 (Least deprived) | 473 (10.2) | 448 (9.3) |  |
| **<u>Oral hygiene practices</u>** |  |  |  |
| Frequency of brushing teeth |  |  | 0.001 |
| Twice a day or more | 1652 (80.8) | 1765 (85.1) |  |
| Less than twice a day | 393 (19.2) | 310 (14.9) |  |
| Used dental floss in the last year = No | 1000 (80.6) | 1013 (76.1) | 0.006 |
| Pattern of dental attendance = Only when<br>in trouble/never | 428 (18.3) | 432 (17.1) | 0.28 |
| **<u>Orthodontic treatment experience</u>** |  |  |  |
| Ever had bracket fitted or adjusted = No | 1493 (88.9) | 1446 (83.1) | <0.001 |
| **<u>Dietary habits</u>** |  |  |  |
| Frequency of eating cakes or biscuits |  |  | 0.018 |
| Twice or more a day | 1040 (44.8) | 1024 (40.5) |  |
| Once or less a day | 1280 (55.2) | 1506 (59.5) |  |
| Frequency of eating sweets |  |  | 0.586 |
| Twice or more a day | 966 (41.6) | 1074 (42.4) |  |
| Once or less a day | 1355 (58.4) | 1460 (57.6) |  |
| Frequency of drinking soft drinks containing sugar |  |  | <0.001 |
| Twice or more a day | 854 (37.1) | 845 (33.5) |  |
| Once or less a day | 1451 (62.9) | 1674 (66.5) |  |
| Frequency of drinking energy drinks |  |  | <0.001 |
| Twice or more a day | 416 (18.1) | 354 (14.2) |  |
| Once or less a day | 1886 (81.9) | 2141 (85.8) |  |
| Frequency of drinking fizzy drinks |  |  | <0.001 |
| Twice or more a day | 1103 (47.3) | 1062 (41.8) |  |
| Once or less a day | 1225 (52.7) | 1476 (58.2) |  |
| Frequency of drinking water |  |  | 0.049 |
| Twice or more a day | 1631 (70.7) | 1810 (71.9) |  |
| Once or less a day | 675 (29.3) | 706 (28.1) |  |
| **<u>Dental anxiety</u>** |  |  |  |
| Self-rated dental anxiety score |  |  | <0.001 |
| Low/no anxiety | 935 (40.9) | 576 (23.4) |  |
| Moderate anxiety | 1172 (51.2) | 1461 (59.5) |  |
| Extreme anxiety | 181 (7.9) | 420 (17.1) |  |
| **<u>Dental health outcomes</u>** |  |  |  |
| d<sub>3</sub>mft (mean (SD)) | 0.8 (1.8) | 0.7 (1.6) | <0.001 |
| D<sub>3</sub>MFT (mean (SD)) | 0.9 (1.9) | 1.1 (2.2) | <0.001 |
| d<sub>1</sub>mft (mean (SD)) | 1.2 (2.2) | 1.0 (2.1) | 0.001 |
| D<sub>1</sub>MFT (mean (SD)) | 1.7 (3.0) | 1.9 (3.1) | 0.012 |
| **<u>BPE (N %)</u>** |  |  |  |
| 0 (N %) | 530 (49.8) | 581 (51.4) | >0.05 |
| ≠ 0 (N %) | 534 (50.2) | 550 (48.6) | >0.05 |

**Note:** Data are presented as frequency (%) unless specified. Numbers may not sum to totals due to missing values; percentage may not sum to 100 due to rounding.
Deprivation index was grouped in quintiles as follows:
1 (most deprived, ranks 1-6496), 2 (ranks 6497-12993), 3 (ranks 12994-19489), 4 (ranks 19490-25986), 5 (least deprived, ranks 25987-32482)
P-value was for comparing characteristics between male and female.
SD: standard deviation

For dental caries comparison between male and females, no differences were found in 5- and 8-year-old children at either caries threshold. Among 12-year-old children, females were associated with higher DMFT scores, regardless of the threshold at which dental caries was diagnosed [IRR (D3): 1.28, 95%CI (1.10-1.49), p<0.01 and IRR(D1): 1.22, 95% CI (1.10-1.35), p<0.001]. The discrepancy in caries experience between genders was evident among 15-year-old children too, but only at the dentinal threshold [IRR (D1): 1.16, 95% CI (1.00-1.35), p<0.05]. Also, the possibility for 15-year-old females to be “caries-free” (DMFT:0) was lower compared to males [OR(D3):0.59, 95% CI (0.45-0.82), p<0.01 and OR(D1):0.55, 95% (0.41-0.74), p<0.001], regardless of threshold at which dental caries was examined. However, when caries experience was estimated including early enamel lesions (D_1_MFT), males matched females in caries experience (Table 4).

**Table 4.**
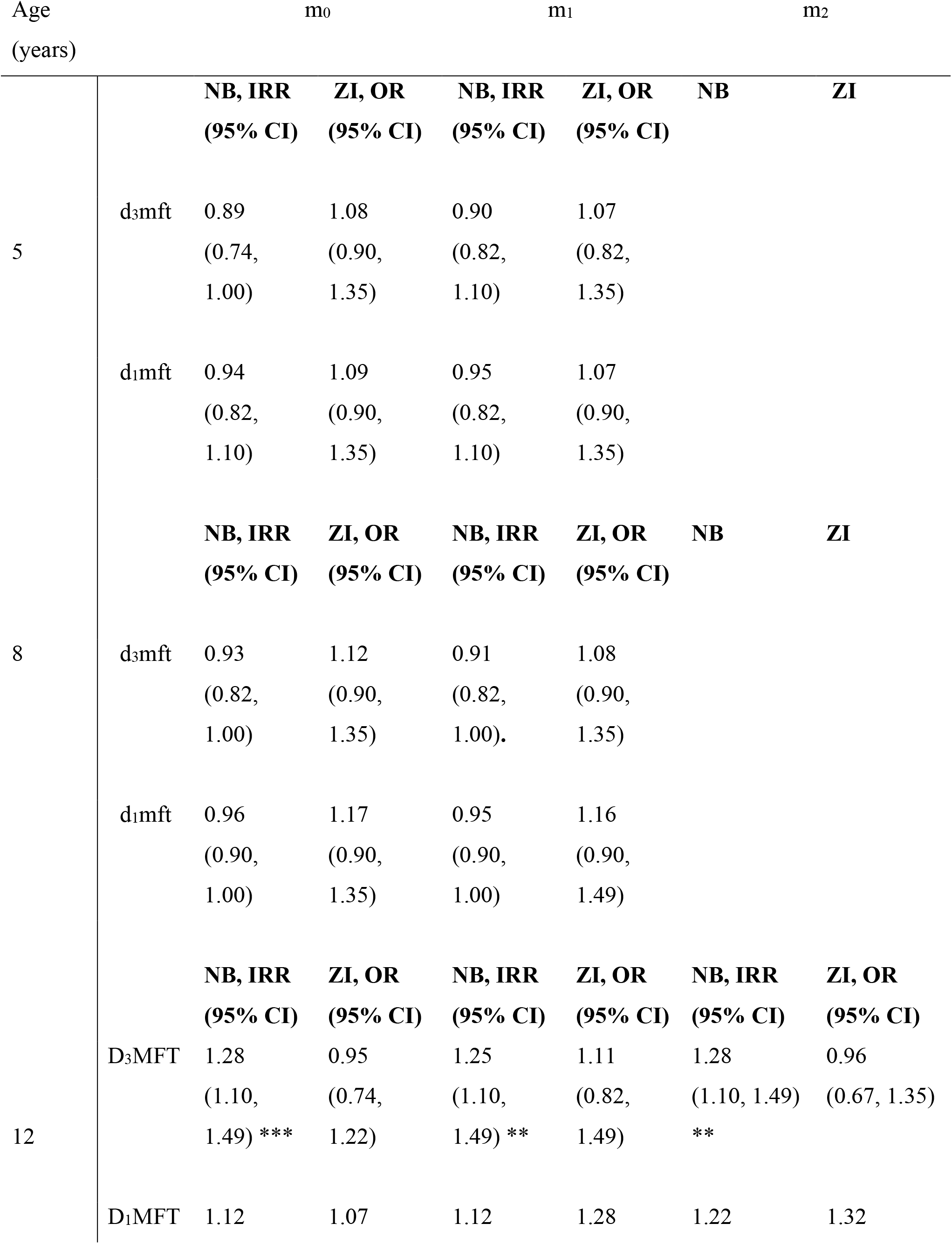
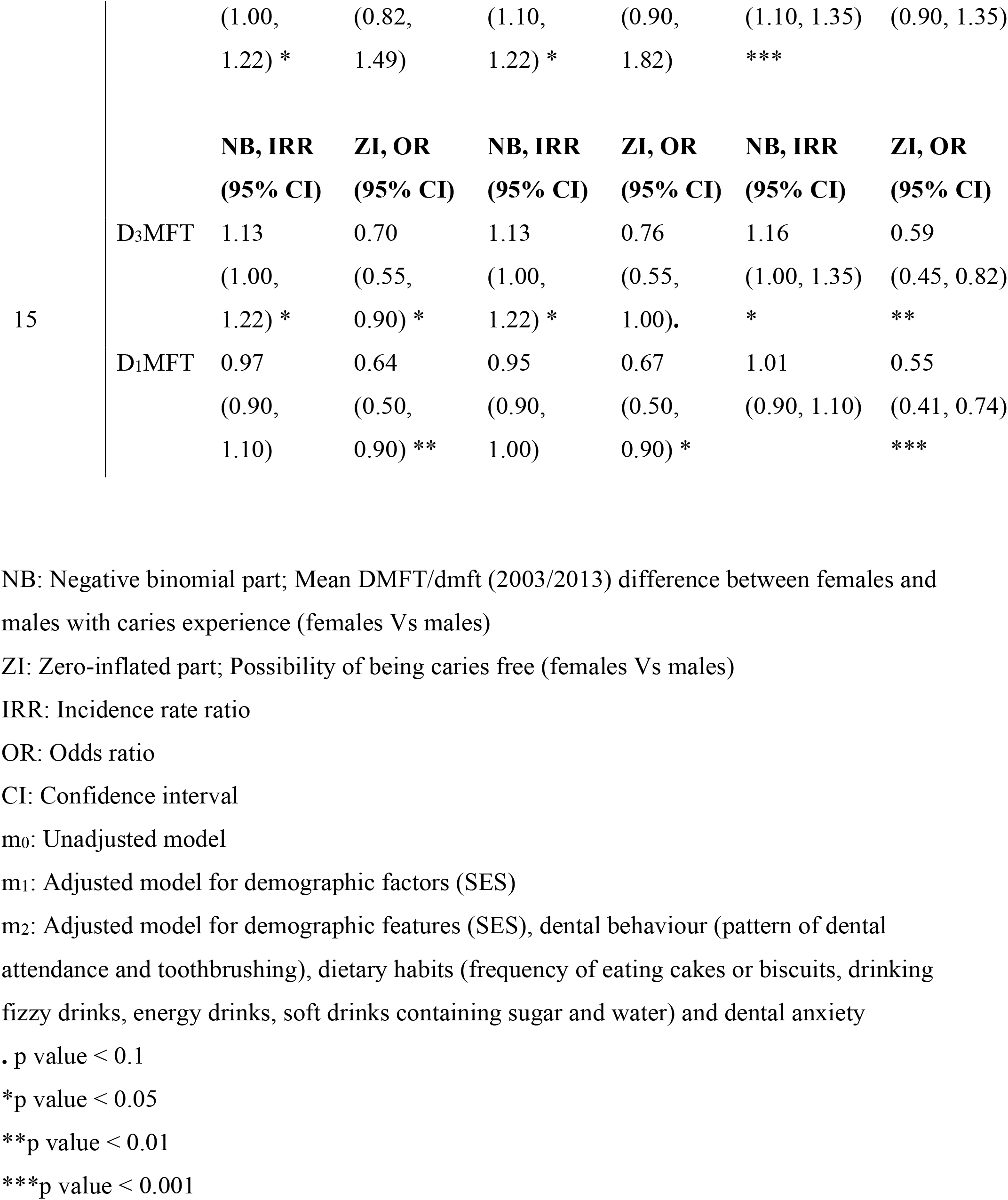
Dental caries experience between females and males (females vs males).

The results of the MLR model are summarised in Table 5, comparing the periodontal status of the 15-year-old females and males. No significant difference (p>0.05), for any of the examined BPE scores, was noted between genders.

**Table 5.**
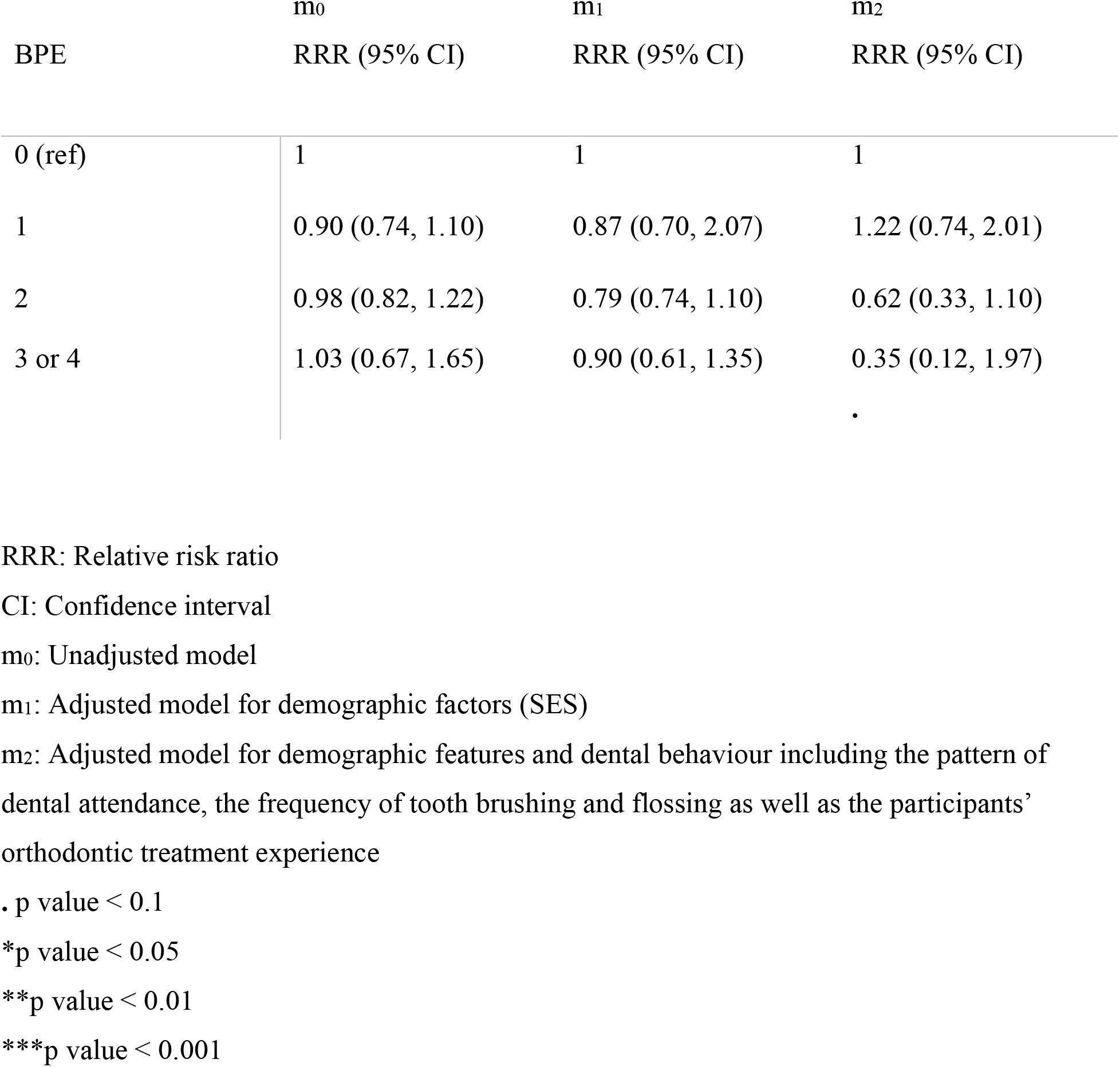
Periodontal status between the 15-year-old females and males (females Vs males).

## Discussion

No gender inequalities in either disease was noted at the primary dentition stages. A potential explanation of the absence of caries inequalities could be that children of such young age cannot opt for their dietary habits nor are responsible for their daily oral hygiene. Therefore, the most important risk factors are influenced directly by parents or guardians, which could demarcate the absence of gender susceptibility to dental caries. Notably, the majority of studies are in line with the lack of significant gender discrepancy regarding dental caries experience in the primary dentition^13,16,32,33^. However, it would be prudent not to draw such conclusion due to lack of data on eating habits and oral hygiene practices for these age groups.

For the permanent dentition, the outcomes were substantial. Females in the 12-year-old group, had higher D_1_MFT and D_3_MFT scores. This susceptibility of females was confirmed among the 15-year-old adolescents too, but only when disease was diagnosed into dentine (D_3_MFT). When early enamel lesions were included in caries diagnosis, the 15-year-old males matched females in caries experience, which denotes that the caries experience of males comprised of more enamel lesions than females.

According to pupil questionnaires, female adolescents consumed sweets and sugary soft drinks less frequently, while they used to brush their teeth more frequently. Ideally, this should have materialised in less exposure to caries. However, higher prevalence of caries among females may be associated with erroneous assumptions on healthy and balanced diet from a dental perspective. It could be hypothesised that participants may have underestimated the daily consumption of hidden sugars with exponential cariogenic risk^34-36^. Another explanation of the higher female susceptibility may be associated with the earlier tooth eruption and, therefore, longer exposure to cariogenic products in the oral cavity^37^. Also, a number of genome-wide-association-studies (GWAS), support that gender differences may be attributed to a number of gene variations influencing the oral environment, enamel formation, dietary preferences and composition of pathogenic bacteria^1,38,39^. Furthermore, differences in saliva composition and in overall saliva flow rate could stem from hormonal changes due to puberty onset^40^, pregnancy or menstruation, rendering female adolescents more prone to dental caries^10,41^. Among physiological variations between genders, elevated oestrogen levels in females have been associated with changes in saliva composition and reduction in the salivary flow. These conditions in combination with a cariogenic diet could contribute to caries incidence among female adolescents^42^.

Although there is no consensus regarding the existence of specific gender inequalities in caries experience for the permanent dentition of adolescents, a number of studies^3,5,8,43^ were in accordance with our findings.

Multiple studies^22,23,44^ were in agreement with the absence of gender discrepancies with regard to the periodontal status of the 15-year-old adolescents based on BPE scores. In contrast, other studies^9,10,13,14^ reported that males had worse gingival or periodontal status than females. The reported susceptibility of male adolescents to poorer periodontal/gingival status, may stem from suboptimal oral hygiene regime rather than the genetic superiority of females. Also, notable deviations in study design, sample selection and indexes used do not allow comparisons or concrete conclusions to be drawn.

The high-quality dataset derived from the 2013 CDHS offers the opportunity for comprehensive analysis of children’s oral health^15^ and serves as a strength of this study. Also, the different thresholds at which dental caries diagnosis was estimated are reflected eloquently in the present study, eliminating the risk of confusion or result deviations and allowing comparisons between different outcomes. Another strength refers to the ZIBN model used to investigate caries experience, which is the most reliable and appropriate statistical approach according to DMFT/dmft distribution^45^.

There are also limitations. A number of factors may mean that CDHS underestimates the true population levels of oral disease. As with most surveys, CDHS was based solely on clinical examination, and epidemiological convention calibrates examiners to “underdiagnose” the presence of dental caries in case of uncertain diagnosis^15^. Also, the BPE index may underestimate periodontal disease^46^. Additionally, children attending special schools, where oral disease levels may be higher, were not included in the CDHS which may have underestimated the burden of such oral diseases among British children^47,48^. A further limitation is that self-report of dietary habits for the 12- and 15-year-old groups may have underestimated the consumption of cariogenic food and drinks. The dietary data being limited to these age groups also means that the ZINB model was not adjusted for those factors for the 5- or 8-year-old groups, which may have rendered the estimations of the d_3_mft and d_1_mft more susceptible to errors. Furthermore, BPE data were only available for the 15-year-old group, rendering comparisons between age groups impossible.

In this study, the first to report on gender inequalities using a large UK dataset which includes both enamel and dentine caries, it is notable that female adolescents were more susceptible to dental caries, when disease examined into dentine (D_3_MFT). When early enamel lesions included in dental caries diagnosis (D_1_MFT), 15-year-old males matched females in their caries experience. These findings suggest that caries experience of males comprised of more enamel lesions and therefore, gender discrepancy at younger ages or when caries examined into dentine only, may be contributed to the earlier tooth eruption in girls. With regard to periodontal status, no gender inequalities were noted among British adolescents.

Despite the current findings, more prospective studies would be required to ascertain accurate conclusions, before stricter preventative measures are recommended for either of genders. Additionally, further investigation on the possible impact of the individual regional water fluoridation scheme may have in children’s caries experience, would be useful. This investigation could probably reflect the need for further preventative measures, like the artificial water fluoridation in more areas across the UK. With respect to children’s periodontal status, more prospective studies along with the usage of a widely agreed index, would be useful in investigating the potential for advanced periodontal treatment needs among children of different ages.

## Data Availability

The Children Dental health Survey 2013 data is public accessible and free to download to all registered users in the UK Data Service: https://beta.ukdataservice.ac.uk/datacatalogue/studies/study?id=7774&type=Data%20catalogue#!/access-data

https://digital.nhs.uk/data-and-information/publications/statistical/children-s-dental-health-survey/child-dental-health-survey-2013-england-wales-and-northern-ireland

## Acknowledgements

We would also like to thank Ioannis Pilalas for his contribution in this publication.

